# Stakeholder perspectives on the potential introduction of a novel tuberculosis vaccine for adolescents and adults in Pakistan: A qualitative study

**DOI:** 10.64898/2026.08.26.26360963

**Authors:** Zoyia Hassan, Zoya Zurez, Muhammad Saad, Nazia Ahsan, Rebecca A Clark, Richard G White, Momin Kazi, Kristin N. Nelson

## Abstract

**Background:** Tuberculosis (TB) remains a major public health challenge globally, with Pakistan ranking among the highest TB burden countries worldwide. Although several novel TB vaccine candidates for adolescents and adults are advancing through late-stage clinical trials, little is known about how these vaccines may be introduced in high-burden settings such as Pakistan. Understanding stakeholder perspectives is crucial for informing early implementation planning and policy development.

**Methods:** We conducted an exploratory qualitative study using semi-structured in-depth interviews with key stakeholders involved in TB control, immunization, clinical care, and health policy in Pakistan. Participants were purposively selected from national and provincial TB programs, Expanded Programme on Immunization (EPI), clinical settings, and academia. Interviews were conducted in English or Urdu, audio-recorded, transcribed verbatim, and analyzed using reflexive thematic analysis following Braun and Clarke’s framework. A hybrid deductive– inductive coding approach was used.

**Results:** Ten stakeholders participated including one whose interview also served as a pilot test of the interview guide. Participants expressed strong support for the introduction of a new TB vaccine, driven largely by Pakistan’s high TB burden and the limitations of current prevention strategies. However, support was based on the availability of strong evidence regarding vaccine safety, effectiveness, and feasibility. Key barriers to vaccine acceptability included low perceived risk of TB, misinformation, stigma, sociocultural influences, and limited public awareness. Stakeholders emphasized community engagement, trusted healthcare providers, and effective communication as critical enablers. Health system challenges included workforce shortages, cold chain limitations, and financing constraints. Household contacts of TB patients were consistently identified as the priority group followed by adolescents and people living with HIV. A phased implementation strategy was broadly preferred followed by gradual integration into existing health services.

**Conclusion:** Stakeholders in Pakistan broadly support new TB vaccines for adolescents and adults. Successful implementation will require addressing sociocultural barriers, strengthening health system capacity, and developing context-specific delivery and prioritization strategies. Early stakeholder engagement and implementation planning are essential for meaningful public health impact in Pakistan.

## Introduction

Tuberculosis (TB) remains the leading cause of death from a single infectious agent globally. According to the World Health Organization (WHO) Global TB Report 2025, there were an estimated 10.7 million cases and 1.3 million deaths attributable to TB in 2024 [1]. Despite decades of control efforts, progress towards achieving global TB elimination targets remain slow. Pakistan, ranked fifth among high-burden countries, bears 73% of the TB burden in the Eastern Mediterranean Region, with an estimated 686,000 incident TB cases and 47,000 deaths annually [1]. This is compounded by socioeconomic vulnerability, health system constraints, and persistent public health challenges within communities [2].

The Bacillus Calmette–Guérin (BCG) vaccine provides protection against extrapulmonary TB in young children but offers limited protection against pulmonary TB in adolescents and adults, the populations most responsible for transmission of *Mycobacterium tuberculosis* and the age groups with the greatest burden of disease [3]. Several new TB vaccine candidates are currently in late-stage clinical trials [4,5]. The most advanced candidate is M72/AS01_E_, which demonstrated approximately 49.7% (90% CI 12.1 to 71.2) efficacy in preventing progression to active TB disease in adults who were IGRA-positive at the time of vaccination [6]. Other candidates, such as MTBVAC [7] and VPM1002 [8], are in late-stage clinical trials.

Efficacy alone does not guarantee successful introduction of a new vaccine [9]. TB vaccines require careful consideration of aspects such as health system readiness, sociocultural context, and public trust. In LMICs like Pakistan, these challenges are compounded by constrained resources, fragmented healthcare systems, and competing public health priorities [10]. The need for careful planning is especially important for TB vaccines for several reasons. First, unlike childhood vaccines delivered through established Expanded Programme on Immunization (EPI) systems, adult and adolescent vaccination will require new or adapted delivery strategies [11]. Second, TB-related stigma can reduce motivation for preventive interventions [12]. Third, early vaccine supply may be limited, requiring decisions about prioritizing high-risk populations [13].

In this context, qualitative research is essential to understand how key stakeholders perceive the introduction of new TB vaccines and to identify potential barriers and facilitators to implementation [14]. Similar stakeholder-focused studies have recently been conducted in other high-burden settings, including South Africa [15] and Indonesia [16], and though these studies have shown that stakeholder perceptions of new TB vaccines vary by setting, none have focused on Pakistan. Stakeholders including policymakers, program managers, clinicians, and researchers play a central role in shaping vaccine introduction strategies. This study therefore explores stakeholder perspectives on the potential introduction of new TB vaccines for adolescents and adults in Pakistan. By examining acceptability, delivery feasibility, prioritization strategies, and anticipated uptake, the study aims to generate policy-relevant insights that can inform early planning for TB vaccine introduction in Pakistan.

## Methods

### Study Design and Setting

We conducted an exploratory qualitative study using semi-structured in-depth interviews in Pakistan, a high TB-burden country with a decentralized health system. TB control activities are coordinated through the National TB Control Program in collaboration with provincial TB programs, while immunization services are delivered through EPI. Vaccine introduction decisions require coordination across federal, provincial, and district-level authorities.

### Participant Selection and Recruitment

Participants were selected using purposive sampling to ensure representation across TB control, immunization planning, clinical care, and health policy. Eligible participants included national and provincial TB program personnel, EPI officials, clinicians involved in TB diagnosis and treatment, and public health researchers and policy advisors. Participants were identified through professional networks and institutional affiliations. Invitations were sent via email or direct communication. Sampling aimed to capture diverse perspectives across policy, programmatic, and clinical domains. Recruitment continued until thematic saturation was reached, which was possible since transcripts from each interview were analyzed soon after they were conducted. Saturation was assessed iteratively by the two coding researchers through concurrent data collection and analysis, whereby transcripts were reviewed and coded after each interview. Saturation was considered achieved when interviews yielded no new codes or themes. All stakeholders who were approached agreed to participate. No one refused or withdrew from the study.

### Data Collection

Interviews were conducted between November 2024 to April 2025 in English and Urdu depending on participants preference, lasted approximately 45-60 minutes and were conducted in person or virtually using a secure online platform. No third parties were present during interviews beyond the interviewer and the participant. One participant was interview twice, once as part of the interview guide and once as a substantive study interview. All other participants were interviewed once only. An interview guide was developed based on the study objectives and relevant literature on vaccine introduction and TB control exploring perceptions of vaccine need, acceptability and barriers, delivery pathways, target populations prioritization, health system readiness, and expected uptake (S1 Text). The guide was pilot tested with one participant prior to commencement of the study. Based on feedback, minor refinements were made to the wording and sequencing of questions before data collection began. As the changes were minor and did not alter the objectives of the guide, data from the pilot interview were retained and included in the final analysis. With participant consent, interviews were audio-recorded and subsequently transcribed verbatim. Urdu interviews were translated into English for analysis. Field notes were maintained throughout data collection to capture contextual observations, emerging themes and served as a supplementary tool for ongoing saturation assessment.

### Data Analysis

Data was analyzed using reflexive thematic analysis as described by Braun and Clarke [17], following six phases: familiarization with the data, generation of initial codes, identification of candidate themes, review and refinement of themes, definition and naming, and production of the analytic narrative. A hybrid deductive–inductive coding approach was employed. Deductive codes were informed by the study objectives and interview guide domains, including acceptability, delivery strategies, prioritization, and system readiness. Inductive codes were generated from participant narratives. Two researchers independently reviewed and coded transcripts iteratively with regular discussions to resolve discrepancies and refine the thematic framework. Coding was managed in NVivo 13. Reflexivity, defined as the ongoing critical examination of how researchers’ own backgrounds, assumptions and personality may shape data interpretation and theme development, was incorporated through team discussions and iterative coding review and reflective memos writing throughout analysis.

### Ethical Considerations

Ethical approval was obtained from the institutional review boards of Aga Khan University (IRB reference #: 2025-10755-32849) and Emory University (IRB reference #: STUDY00003098). All participants provided informed consent prior to participation. Identifying information was removed from transcripts and data was stored in password protected files accessible only to the research team.

## Results

Ten in-depth interviews were conducted with stakeholders including one interview that also served as a pilot test of the interview guide. It included respondents from the Ministry of Health (n=2), National TB Program (n=3), TB Clinicians (n=2), TB researchers (n=1), and Immunization program personnel (n=2). Respondents’ perspectives across policy, program implementation, clinical and research domains. The distribution of participants across stakeholder groups summarised in the Table 1.

**Table 1.** Distribution of participants across stakeholder groups.

| Stakeholder Category | Participants (n) | Description of Roles |
| --- | --- | --- |
| Ministry of Health | 2 | Policy and strategic leadership |
| National TB Program Personnel | 3 | Program implementation and management |
| TB Researchers | 1 | Academic and research expertise |
| TB Clinicians | 2 | Clinical care and patient management |
| Immunization Program Personnel | 2 | Vaccine delivery and program planning |
| **Total** | **10** |  |

### Interest and Support for New TB Vaccine

Stakeholders across all groups expressed strong interest in the introduction of a new TB vaccine, driven primarily by recognition of Pakistan’s high TB burden and the limitations of existing prevention strategies. Participants consistently viewed a new vaccine as a necessary addition to current efforts, particularly given ongoing transmission despite widespread availability of treatment. Many described the vaccine as a potentially transformative intervention:

> *“Given the high TB burden and the socio-economic conditions in our country, this could be a real gamechanger and a cost-effective intervention.” (Immunization program personnel)*

Despite strong enthusiasm, participants highlighted that policy decisions would depend on clear and credible evidence on vaccine safety, effectiveness, and feasibility. There was a strong preference for locally generated or contextually relevant evidence reflecting concern that global recommendations may not resonate with national decision makers or the public:

> *“There is interest and excitement about a new TB vaccine, but we cannot move forward without strong evidence.” (National TB Program personnel)*
>
> *“We cannot introduce a TB vaccine just based on global recommendations. People here will ask whether it has worked in countries like India, Bangladesh, or sub-saharan Africa where the health system and TB burden are similar to ours". (National TB Program personnel)*

### Barriers to Acceptability

Misinformation and persistent TB-related stigma were the most prominent barriers to uptake. The association of TB with poverty and social exclusion was expected to discourage community engagement with preventive interventions. Trust in vaccination was described as strongly influenced by community leaders, elders, and religious figures:

> *“Acceptance is strongly influenced by community leaders, elders, and religious figures. Without their support, uptake will remain low.” (Immunization program personnel)*

Low awareness of TB prevention measures and the unfamiliar concept of vaccination for TB, a disease typically associated with long drug regimens rather than prevention, were additional barriers. Existing communication channels were described as largely focused on childhood immunization, with few strategies targeting adults:

> *“We do not have strong or structured communication channels for adult vaccination, so correct information does not reach people effectively.” (Immunization program personnel)*

Geographic inequities further compounded access challenges in remote and underserved areas, where even willing individuals may face insurmountable logistical barriers:

> *“In many remote areas, services are simply not available. Even if people are willing, access becomes the main barrier.” (TB Researcher)*

### Strategies to Improve Acceptability

Early community engagement and social mobilization were identified as essential prerequisites. Trusted health care providers and community leaders were seen as key messengers, capable of addressing concerns and building confidence before rollout. COVID-19 communication strategies were widely cited as an effective model:

> *“We need COVID-style messaging through TV, newspapers, and digital media to reach the general population and normalize the TB vaccine.” (TB program stakeholder)*
>
> *“Before any vaccination campaign, community meetings are essential. When local influencers and caregivers are involved early, acceptance improves.” (Provincial TB program stakeholder)*

A specific concern emerged regarding perceived vaccine origin: vaccines seen as foreign-funded may face politically motivated hesitancy in some communities in Pakistan, making transparent communication about development and sourcing a priority:

> *“People here often question where the vaccine is coming from. If they hear it is funded by America or developed with support from India, some communities may become hesitant because of political beliefs and mistrust. Clear communication about who developed the vaccine, how it was tested, and whether it is safe for our population will be very important.” (TB Researcher)*

### Health System Readiness and Logistical Feasibility

Stakeholders raised substantial concerns about health system preparedness for adult vaccination, which falls outside routine service delivery mechanisms. Workforce shortages, cold chain limitations, and financing gaps were recurring themes:

> *“Adult vaccination is not part of the routine service flow. Clinics are already overcrowded, and adding another vaccine will disrupt daily operations.” (Provincial TB program stakeholder)*
>
> *“Without dedicated funding, it will be difficult to manage procurement, staffing, and expansion of cold chain for a new vaccine.” (Policy-level stakeholder)*

Both EPI and the TB program were seen as potential delivery platforms, each with important limitations. EPI’s established infrastructure was seen as a feasible foundation but would require significant adaptation to include adult vaccination. The TB program was valued for its ability to identify high-risk populations but existing staff are already overburdened:

> *“EPI has experience with vaccines, but adult TB vaccination is outside its usual mandate and would need major system adjustments.” (Immunization program personnel)*
>
> *“The TB program knows the high-risk populations, but staff are already overburdened with diagnosis and treatment responsibilities.” (TB program personnel)*

Participants broadly favored a coordinated, multi-platform approach rather than reliance on any single system.

### Target Population Prioritization and Implementation Strategies

Stakeholder responses demonstrated clear convergence on priority groups for TB vaccination (Table 2). Household contacts of TB patients emerged as the most consistently prioritized group (n=4; 16%), reflecting strong agreement on their elevated risk and programmatic accessibility through existing contact tracing:

> *“If we want maximum impact, household contacts of TB patients should be the first group to receive the vaccine.” (Provincial TB program stakeholder)*.

**Table 2.** Priority target groups for TB vaccination among adults and adolescents in Pakistan: frequency of stakeholder mentions and participant affiliations.

| Serial No: | Target Group | Mentions (N)<br>(%) | Participant affiliations<br>identifying this target<br>groups |
| --- | --- | --- | --- |
| 1 | Household contacts of TB<br>patients | 4 (16) | TB program personnel,<br>Ministry of Health |
| 2 | Adolescents | 3 (12) | TB program Personnel |
| 3 | HIV-positive individuals | 3 (12) | TB Program Personnel |
| 4 | nmunocompromised individuals | 2 (8) | TB program Personnel, TB Clinician |
| 5 | Low SES / Slums | 2 (8) | Immunization and TB personnel |
| 6 | Older adults | 2 (8) | TB program Personnel |
| 7 | Women of reproductive age | 2 (8) | TB program Personnel |
| 8 | Young adults | 2 (8) | Ministry of health, TB Program |
| 9 | Healthcare workers | 2 (8) | Immunization program Personnel |
| 10 | Children (under five years of age) | 1 (4) | TB Clinician |
| 11 | Immigrants | 1 (4) | TB program Personnel |
| 12 | Prisoners | 1 (4) | TB Program Personnel |
| 13 | Total | 25 (100) |  |

Adolescents and people living with HIV (each n=3; 12%) were also high-priority, indicating recognition of both epidemiological risk and programmatic importance. Several other groups were moderately prioritized (each n=2; 8%), including immunocompromised individuals, populations living in low socioeconomic settings or slums, older adults, women of reproductive age, children, and healthcare workers.

Stakeholders consistently recommended a phased approach to vaccine introduction, beginning with pilot programs in high-burden areas followed by mass campaigns targeting adults and adolescents outside routine systems, with eventual integration into existing service delivery platforms:

> *“We need to start with pilots, then use mass campaigns to reach adults, and only later integrate the vaccine into routine services." (Immunization Program Stakeholder)*

The COVID-19 vaccine rollout was viewed as both a model and a caution demonstrating what is operationally possible under strong political will, while underscoring that such emergency-level mobilization is unlikely to be replicated for TB:

> *“COVID was treated as an emergency with unlimited resources. TB will not receive that level of attention.” (TB program personnel)*.

Regarding uptake expectations, stakeholders anticipated modest initial coverage with gradual improvement as trust builds. TB vaccine uptake was expected to exceed that of TB preventive therapy (TPT) given the relative simplicity of vaccination, but fall short of COVID-19 uptake due to lower perceived urgency:

> *“In the beginning, uptake will be slow, but as people see others getting vaccinated safely, acceptance will improve over time.” (TB program personnel)*.

Similarly, another participant highlighted that

> *“Vaccine is easier than months of preventive treatment, so acceptance is expected to be higher than TPT.”* (TB clinician).

## Discussion

This study provides stakeholder insights on TB vaccine introduction in Pakistan, one of the highest-burden settings globally. Overall, stakeholders expressed strong support for a new TB vaccine, but this support was based on evidence of safety, effectiveness, feasibility of integration into local health systems, and efforts to engage key community leaders to ensure broad acceptance. Key barriers to acceptability included low perceived risk of TB, misinformation, stigma, and limited public awareness, while community engagement and trusted healthcare providers were viewed as critical enablers. Health system readiness was a further concern, particularly workforce shortages, cold chain limitations, and financing constraints. Household contacts of TB patients were the most consistently prioritized target group, followed by adolescents and people living with HIV, and a phased implementation strategy beginning with pilot programs was broadly preferred.

Our findings are consistent with emerging evidence from high-burden settings about attitudes towards new TB vaccines. For example, a recent study in Mozambique found generally high willingness to accept new TB vaccines among adults, adolescents, and caregivers, but similarly found that acceptance depended on trust, perceived safety, and clear information about vaccine benefits. [18] Likewise, qualitative research on new TB vaccine implementation in South Africa has highlighted cautious optimism among stakeholders, coupled with the need for strong evidence and implementation planning [15]. The call for "strong evidence" is notable. The most advanced TB vaccine candidate, M72/AS01_E_, demonstrated 49% (90% CI 12.1 to 71.2) efficacy against active TB disease in a Phase 2b trial [6]. While this efficacy is only moderate, modelling suggests that even vaccine with as low as 20% efficacy could avert millions of TB cases and deaths when deployed at scale in high burden settings[19,20] Stakeholder expectations around efficacy will therefore need careful management through transparent communication, helping decision-makers and communities understand that even a moderately effective vaccine, combined with existing TB control measures, may be sufficient where the unmet need is large.

The barriers to new TB vaccine introduction that were identified such as stigma, misinformation, and low awareness are consistent with both TB-specific [21] and broader vaccine literature [22,23]. The intersecting nature of these barriers is particularly important in Pakistan: TB-related stigma, which links the disease to poverty and social exclusion, may uniquely deter individuals from seeking vaccination in ways not observed for other vaccines. The influential role of community and religious leaders in shaping health behaviors is well-documented in LMIC settings [24], and in Pakistan, where community networks are central to decision-making, their early engagement will be essential. A uniquely Pakistan-specific finding was concern around perceived vaccine origin and the possibility that vaccines seen as foreign-funded may face politically motivated hesitancy. This finding highlights the need for proactive, transparent communication tailored to local sensitivities.

Priority populations identified in Pakistan included household contacts, adolescents, and PLHIV, which closely mirror those identified in South Africa and Indonesia [15,16]. However, the practical reachability of these groups varies in Pakistan’s context. Household contact tracing remains inconsistent across provinces, meaning systematic identification of this group would first require programmatic strengthening. Adolescents are largely reachable through schools, and Pakistan’s 2025 GAVI-supported HPV vaccination campaign provides a recent precedent for adolescent delivery through EPI [25]; however, out-of-school adolescents will require separate outreach strategies. PLHIV can be reached through existing HIV care services; however, the feasibility of this approach depends on the proportion of TB cases in Pakistan that occur among PLHIV, which is lower than in southern African settings with high TB-HIV co-burden. [26]. Critically, delivery strategies for adults remain the least defined and most challenging, as no established adult vaccination infrastructure exists in Pakistan, requiring deliberate advance planning and investment well before any vaccine becomes available [27].

Stakeholders’ strong emphasis on locally generated evidence is also consistent with broader vaccine introduction literature. Studies examining vaccine policy decision-making in LMICs have shown that adoption is heavily influenced by context-specific data on burden, feasibility, and cost-effectiveness [13]. In the context of TB vaccines, the absence of local clinical trial data from Pakistan may represent a significant barrier to policy adoption. Current pivotal trials are planned or ongoing in Africa and other countries in Asia (Indonesia, India), but at present no trials are planned in Pakistan. This concern has also been raised in global TB vaccine preparedness frameworks, which emphasize the importance of early planning for local data generation and post-introduction evaluation [28].

Several limitations should be acknowledged. The sample size of ten participants, while appropriate for purposive expert sampling in a novel policy area, may not fully capture all relevant stakeholder perspectives. Financing stakeholders were not included, representing a gap given the centrality of cost considerations for any new vaccine program. Community members and patients were also not included; their perspectives may differ meaningfully from those of institutional stakeholders. Findings reflect anticipated rather than real-world implementation experiences and may evolve as evidence on vaccine performance, cost, and delivery strategies emerges.

In conclusion, stakeholders in Pakistan are broadly supportive of new TB vaccines for adolescents and adults. This study adds important context-specific insights to a growing multi-country evidence base, particularly regarding the role of local evidence, the influence of perceived vaccine origin on trust, and the challenges of adult vaccine delivery in a decentralized health system. Successful introduction will require advance planning that addresses sociocultural barriers, strengthens health system capacity, and develops context-specific delivery and prioritization strategies, supported by tailored communication and sustained early engagement with communities and decision-makers.

## Supporting information

Interview guide

## Data Availability Statement

All relevant data supporting the findings of this study are presented within the manuscript and its Supporting Information files. Interview transcripts cannot be made publicly available to protect participant confidentiality, as disclosure could risk re-identification of participants. Further, participants have not provided informed consent for their data to be stored in a public repository. Requests for access to the underlying data may be directed to the corresponding author at

## Author Contributions

**Conceptualization:** KN, MK. **Methodology:** NA, ZH, RAC, RGW. **Software:** ZH. **Validation**: ZH, ZZ. **Formal Analysis**: ZH **Resources**: KN, MK, **Data Curation**: ZH, ZZ, MS.

**Writing – Original Draft Preparation:** ZH

**Writing – Review & Editing**: KN, MK, RAC, RGW, ZH. **Visualization**: ZH

**Supervision:** KN, MK, NA. **Project Administration**: ZH, ZZ, MS, NA. **Funding Acquisition**: KN

## Supporting Information

**S1 Text. Interview Guide for the Participants**

