## Supplementary material for "Stakeholder perspectives on the potential introduction of a novel tuberculosis vaccine for adolescents and adults in Pakistan: A qualitative study": Interview guide

**Assessing the Epidemiologic Impact of TB Vaccines in Adults of Pakistan using Mixed Methods Approach**

Semi-structured Interview Guide

**Contents:**

**Introduction material:** (applicable to all categories of stakeholders)

Introduction

Warm-up questions

Main questions

**Stakeholder-specific questions**:

Group A: National TB program personnel, TB researchers, TB clinicians

Group B: Immunization program personnel

**Opening Statement:**

Thank you for agreeing to an interview today. My name is ___ and I am part of a research team at the Aga Khan University collaborating with Emory University in the United States. We are conducting research in preparation for the introduction of new TB vaccines. We feel that it is very important to collect the perspectives and insights of National **TB program personnel, TB researchers, clinicians, immunization program personnel, and other high-level health officials** about how a TB vaccine might be introduced in Pakistan. During this interview today, we will be discussing your opinions on the potential introduction of an adult and adolescent TB vaccine in Pakistan under several different scenarios. This interview should last around 60 minutes. We are most interested in your personal experiences, opinions, and views on the topics that we discuss, so please don’t feel shy. Your views are very valuable to us, and we are here to learn from you.

Before we begin, we would like to state that this interview is entirely voluntary, and if you would like to stop the interview at any time, please let us know. For data collection purposes, and to ensure we collect your complete and accurate perspective, we would like your permission to record our discussion. Any data collected during this interview will remain completely confidential and will only be used by members of the team for the purposes of our study. If at any point during the interview you would like to stop the recording, please let us know and we will adjust accordingly.

Before we begin, do you have any questions or concerns you would like to address?

If at any point you have a question or concern, we will pause and address the issue before continuing.

**Basic Information:**

Tuberculosis (TB) is a major cause of ill health and was the leading infectious cause of death globally in 2019, with 10 million new cases and 1.5 million deaths occurring disproportionately in low- and middle-income countries. To accelerate progress towards the World Health Organization’s (WHO) ambitious goal to cut global TB deaths by 95% and cases by 90% by 2035, new tools to prevent TB are essential. New and repurposed vaccines against TB hold promise to fill this gap.

Several promising vaccine candidates that aim to prevent TB disease in adolescents and adults are currently in vaccine efficacy (phase II and III) trials. Briefly, trials have shown that vaccinating adolescents and adults against TB can reduce the risk of TB infection and disease by about half. Some vaccines under consideration prevent initial TB infection, like MTBVAC, and others would prevent TB disease among those who are already infected, like M72-AS01E. There are several vaccines in phase III trials, which is the final stage of testing before a vaccine could be approved and licensed for use.

Understanding the potential population-level vaccine impact of new TB vaccine programs can inform the development of strategies for introducing new and repurposed TB vaccines into immunization programs once they are licensed and approved for use. The mathematical models developed as a part of this study will help to understand the implications of potential strategies for the implementation of these vaccines, specific to Pakistan.

With this in mind, our goal is to gain a real-world understanding of how a TB vaccine for adolescents and adults may be integrated into the immunization and TB programs in Pakistan. We will use the insights that you provide today to inform the development of models which will aim to estimate the possible impact of introducing an adult and adolescent TB vaccine in Pakistan.

Before we get started, did you have any questions about the background information that was sent to you on the candidate vaccines MTBVAC and M72-AS01_e_?

The remainder of our conversation will center around the scenarios and characteristics of the potential introduction of an adult and adolescent TB vaccine in Pakistan.

Before we begin, I would like to gather some basic background information that is helpful to us:

| Interviewer |  |
| --- | --- |
| Participant code |  |
| Date of interview |  |
| Time of interview | Time started: |
|  | Time ended: |
| Which of the following are your areas of experience? (tick all that are relevant) | - Tuberculosis - Vaccine development - Vaccine procurement/supply - Vaccine delivery - Health/programme financing - Regional/local level - National level - Regulatory bodies - Policy maker e.g., NITAG - Other (please specify): __________________ |

Thank you again for your time and participation in our study.

**General questions:**

1. To begin with, I would like to learn about you and your role at [*ORGANIZATION*].

- Could you describe your current responsibilities?
- Could you describe your past responsibilities?
  - In this role and in previous roles?

Tuberculosis continues to be a critical public health challenge in Pakistan, which accounts for 6.3% of global TB cases, making it one of the high TB-burden countries worldwide. In 2023, Pakistan reported a TB incidence rate of approximately 259 cases per 100,000 population, with a significant proportion of the burden falling on adults, who constitute over 90% of TB cases globally. Existing vaccination strategies, primarily reliant on the BCG vaccine at birth, have shown limited efficacy in reducing TB incidence among adults. New vaccines like M72/AS01E, which prevents progression to active TB in infected individuals, and MTBVAC, designed to prevent initial TB infection, present a unique opportunity to address this challenge. Therefore, we are trying to understand how new TB vaccines might be delivered to adults/adolescents.

2. If an efficacious and safe vaccine were available, do you think there would be interest in a new TB vaccine at the national/regional/individual level?
   a. And would there be any differences between M72/AS01E and MTBVAC?
3. If there were pre-requisites of data you consider critical to your country/program’s ability to make decisions on TB vaccine introduction, what would those be? (e.g., efficacy in specific high-risk group, clinical studies in local context; cost of delivery to adolescents, impact on country TB burden if given to different target groups etc.)

**Potential barriers**

4. What are the characteristics of the vaccine that should be there for its successful delivery? And what is not at all needed in a vaccine for greater acceptability?
5. Are there any potential **logistical** barriers (such as capacity, resources) to introducing an adult/adolescent TB vaccine?
6. Are there any challenges or barriers associated with **integrating a new vaccine with other existing preventative health interventions** among adolescents and adults? (e.g., capacity, resources)
   a. What do you think could be done to help overcome these?
7. Are there any potential **acceptability barriers** at the individual or community level to introducing an adult/adolescent TB vaccine? (e.g., political, importance, religion, safety)
   a. What do you think could be done to help overcome these?
8. What challenges might be faced in introducing **routine** vaccination to adult/adolescents?
   a. What do you think could be done to help overcome these?
9. What challenges might be faced in introducing **mass** vaccination to adult/adolescents?
   a. What do you think could be done to help overcome these?
10. Are there any **other** challenges or barriers to introducing an adult/adolescent vaccine that you think might be important?
    a. What do you think could be done to help overcome these?
11. If those changes could be put into place, what impact do you think they would have on the introduction of the vaccine (e.g., more likely to be introduced, better coverage, better acceptance at the national/community/individual level)?

**Population prioritization and uptake**

Now we will discuss specific populations which may be prioritized for a TB vaccine. First we will focus on the M72-AS01_e_ vaccine candidate.

*Note to interviewer: Tables to help structure the discussion for the subsequent questions are provided at the end of the document*

12. Which populations or groups would you vaccinate (e.g., specific age groups, “high-risk” groups, all individuals)?
13. If there was a limited supply of vaccines, how would the roll out for the groups identified in question 12 be prioritized?

For the target groups identified in question 12, in the order they were prioritised in question 13, please answer:

14. For each of the identified groups, would you use routine vaccination (at a given time or age), or campaigns to vaccinate this group?
15. For each of the identified groups, how and where would the vaccine be delivered as described above? (e.g. during an existing interaction with the health service, at school, at the workplace, or through community campaigns)
16. What vaccination coverage do you think would be achieved in this group (would this be different for routine-maintained coverage, for campaign peak coverage)?
17. For campaigns, what would be the frequency and length of campaigns (e.g., every year, once every three years, etc.)
18. What are some of the challenges or barriers with introducing a new TB vaccine in the identified priority group?
19. For each of the identified groups, what do you anticipate the rough population size is? (1000s, 10 000s,
    100 000s?)
20. Are you aware of any data sources that would indicate the size of these groups and the burden of TB in each group?

Now, we will go through the same questions, but this time for the MTBVAC vaccine candidate.

[REAPEAT QUESTIONS 12-20]

**COVID-19 vaccines**

21. What was the experience in Pakistan with introducing COVID-19 vaccines? (e.g., What went well? What were some of the challenges/barriers with introduction? What was learned that could be used for introducing new vaccines?)
22. Who was it given to? How was it delivered to that population? What coverage was achieved? Or where might we find publicly available data on coverage?
23. Is there any infrastructure remaining from COVID-19 vaccine delivery that could be used to deliver a new adolescent/adult TB vaccine? If yes, what elements are available? What would have to be created?

**Stakeholder specific questions:**

**Group A: National TB program personnel/TB researchers and clinicians**

Next, I'd like to learn about the likely uptake of a TB vaccine compared to other, similar preventive interventions.

As you know, one of the interventions that is currently recommended against TB is TB preventive treatment. According to WHO, 50% of household contacts in Pakistan are placed on TB preventive treatment. (To our knowledge, no data is publicly available on children and people living with HIV, other priority groups for TPT established by WHO.)

1. How would you describe the uptake of TPT in Pakistan?
2. Do you think coverage of an adult TB vaccine would be higher or lower than uptake of TPT? Would it vary across the groups identified in question 12?

**Probe:**

Why do you think coverage of an adult TB vaccine would be higher or lower than uptake of TPT?

3. Are there any challenges or barriers associated with adding a vaccine to the existing TB program? (e.g., capacity, resources)
   a. What do you think could be done to help overcome these?

As you know, the SARS-CoV-2 vaccine was rolled out among adolescents and adults in Pakistan in 2021-22. According to WHO, 60% of the population in Pakistan received at least one dose.

4. How would you describe the uptake of the SARS-CoV-2 vaccine in Pakistan?
5. Do you think coverage of an adult TB vaccine would be higher or lower than the SARS-CoV-2 vaccine? Would it vary across the groups identified in question 12?

**Probe:**

Why do you think coverage of an adult TB vaccine would be higher or lower than the SARS-CoV-2 vaccine?

**Stakeholder specific questions:**

**Group B: Immunization program personnel**

Next, I'd like to learn about the immunization program in Pakistan and vaccination strategies employed for other vaccines.

As you know, the there are several vaccines that are already targeted towards adolescents and/or adults in Pakistan, including tetanus toxoid (TT), HPV and meningococcal vaccine. According to WHO, 94% of the eligible population in Pakistan received at least one dose of the HPV vaccine and 61% of those eligible complete the third dose of TT.

1. How would you describe the current uptake of adult / adolescent vaccines in Pakistan?
2. How are routine adult and/or adolescent vaccines delivered currently?
3. What strategies have been most effective in reaching adults and adolescents with current vaccines?
4. If any of these vaccines were introduced recently (in the past 10 years), what were some of the challenges/barriers to introducing the vaccine(s)?

5. What challenges might arise if a new TB vaccine for adults and adolescents, were added to the existing immunization program? Could these challenges relate to resources, capacity, or infrastructure?

a) Would those same challenges be faced with the introduction of an adult/adolescent TB vaccine?

As you know, the SARS-CoV-2 vaccine was rolled out among adolescents and adults in Pakistan in 2021-22. According to WHO, 60% of the population in Pakistan received at least one dose.

6. How would you describe the uptake of the SARS-CoV-2 vaccine in Pakistan?

7. What strategies were used to deliver SARS-CoV-2 vaccines to adults and adolescents in Pakistan?

8. What strategies were most effective in reaching adults and adolescents with SARS-CoV-2 vaccines?

**Closing Statement:**

We have now completed our interview. We greatly appreciate you taking the time to provide us with your unique insight on our study. Before we end, do you have any final questions that you would like to ask us or anything else you want to share?

Thank you again for participating in this interview, we greatly appreciate your time

**For M72/AS01_E_:**

| Group | a | b | c | d | e | f | g | h | i | j | k |
| --- | --- | --- | --- | --- | --- | --- | --- | --- | --- | --- | --- |
|  | Target population and description | Priority of identified group | Vaccination schedule (routine or campaign) | How and where would the vaccine be delivered? | How long after registration / recommendation would you expect vaccination to start? | Vaccination coverage (routine = maintained, campaigns = total) | How quickly could this coverage be achieved? | *For campaigns:* Frequency/ length of campaigns (years) | Potential challenges/ barriers with introduction | Which stakeholders should be involved? | Population size |
| 1 |  |  |  |  |  |  |  |  |  |  |  |
| 2 |  |  |  |  |  |  |  |  |  |  |  |
| 3 |  |  |  |  |  |  |  |  |  |  |  |

**For MTBVAC:**

|  | a | b | c | d | e | f | g | h | i | j | k |
| --- | --- | --- | --- | --- | --- | --- | --- | --- | --- | --- | --- |
| Group | Target population and description | Priority of identified group | Vaccination schedule (routine or campaign) | How and where would the vaccine be delivered? | How long after registration / recommendation would you expect vaccination to start? | Vaccination coverage (routine = maintained, campaigns = total) | How quickly could this coverage be achieved? | *For campaigns:* Frequency/ length of campaigns (years) | Potential challenges/ barriers with introduction | Which stakeholders should be involved? | Population size |
| 1 |  |  |  |  |  |  |  |  |  |  |  |
| 2 |  |  |  |  |  |  |  |  |  |  |  |
| 3 |  |  |  |  |  |  |  |  |  |  |  |
